# Myocardial Characterization for Early Diagnosis, Treatment Response Monitoring, and Risk Assessment in Systemic Light-Chain Amyloidosis

**DOI:** 10.1101/2023.10.04.23296572

**Authors:** Olivier F. Clerc, Sarah A. M. Cuddy, Michael Jerosch-Herold, Dominik C Benz, Ethan Katznelson, Jocelyn Canseco Neri, Alexandra Taylor, Marie Foley Kijewski, Giada Bianchi, Frederick L. Ruberg, Marcelo F. Di Carli, Ronglih Liao, Raymond Y. Kwong, Rodney H. Falk, Sharmila Dorbala

**Author notes:** Address for Correspondence Sharmila Dorbala, MD, MPH Cardiac Amyloidosis Program, Division of Cardiology, Department of Medicine Cardiovascular Imaging Program, Division of Cardiology and Department of Radiology Division of Nuclear Medicine and Molecular Imaging, Department of Radiology Brigham and Women’s Hospital and Harvard Medical School 75 Francis St., Boston, MA 02115; Twitter/X Handle: @DorbalaSharmila.

## Abstract

**Aims:** In systemic light-chain (AL) amyloidosis, cardiac involvement portends poor prognosis. Using myocardial characteristics on magnetic resonance imaging (MRI), this study aimed to detect early myocardial alterations, to analyze temporal changes with plasma cell therapy, and to predict risk of major adverse cardiac events (MACE) in AL amyloidosis.

**Methods and Results:** Participants with recently diagnosed AL amyloidosis were prospectively enrolled. Presence of AL cardiomyopathy (AL-CMP vs. AL-non-CMP) was determined by abnormal cardiac biomarkers. MRI was performed at baseline and 6 months, with 12-month imaging in AL-CMP cohort. MACE was defined as all-cause death, heart failure hospitalization, or cardiac transplantation. Mayo AL stage was based on troponin T, NT-proBNP, and difference in free light chains. The study cohort included 80 participants (median age 62 years, 58% males). Median left ventricular extracellular volume (ECV) was significantly higher in AL-CMP (53% vs. 30%, p<0.001). ECV was abnormal (>32%) in all AL-CMP and in 47% of AL-non-CMP. ECV tended to increase at 6 months and decreased significantly from 6 to 12 months in AL-CMP (median -3%, p=0.011). ECV was strongly associated with MACE (p<0.001), and improved MACE prediction when added to Mayo AL stage (p=0.002). ECV≤32% identified a cohort without MACE, while ECV>48% identified a cohort with 74% MACE.

**Conclusions:** In AL amyloidosis, ECV detects subclinical cardiomyopathy. ECV tends to increase from baseline to 6 months and decreases significantly from 6 and 12 months of plasma cell therapy in AL-CMP. ECV provides excellent risk stratification and offers additional prognostic performance over Mayo AL stage.

## Introduction

In systemic light-chain (AL) amyloidosis, plasma cell clonal cells produce abnormal immunoglobulin light chains prone to misfolding. These light chains aggregate as amyloid fibrils in multiple organs, expand the extracellular space, lead to organ dysfunction and to death.(1) In the heart, amyloid fibril deposition typically alters myocardial characteristics, cardiac structure and function.(2) Such cardiac involvement is the primary determinant of adverse outcomes in AL amyloidosis.(1) To improve outcomes, several highly effective therapies targeting the plasma cell dyscrasia are currently available for AL amyloidosis.(3) However, the effects of such therapies on myocardial characteristics are not well known.

Magnetic resonance imaging (MRI) is a powerful tool to assess multiple aspects of cardiac disease. In addition to cardiac structure and function, MRI can evaluate myocardial characteristics using late gadolinium enhancement (LGE), T1 and T2 mapping, and can estimate interstitial expansion by the extracellular volume (ECV) fraction.(2) These myocardial characteristics by MRI, are frequently altered in AL amyloidosis and have been associated with mortality.(4–8) Thus, myocardial characteristics, particularly ECV, are emerging as novel markers for myocardial characterization, treatment response monitoring, and outcome prediction in AL amyloidosis.(4–11) However, current knowledge is limited on myocardial characteristics in patients without overt cardiac involvement.(9) It is unclear whether MRI can detect subclinical cardiomyopathy more sensitively than other methods. Furthermore, data on temporal changes in myocardial characteristics during plasma cell therapy are still scarce.(10,11) More evidence is needed to clarify whether AL cardiomyopathy regresses after therapy. Moreover, whether myocardial characteristics can predict adverse outcomes beyond the established, biomarker-based Mayo AL stage is not clearly established.(7,8)

Accordingly, the present study was conducted in participants with recently diagnosed systemic AL amyloidosis with or without cardiomyopathy, and evaluated left ventricular (LV) myocardial characteristics with the following aims: (1) to detect early myocardial changes in participants without overt cardiomyopathy, (2) to quantify temporal changes in myocardial characteristics during plasma cell therapy, and (3) to analyze associations between myocardial characteristics at baseline and adverse outcomes, including their additional predictive value over the Mayo AL stage. We also explored changes in cardiac structure and function and their prognostic value over the Mayo AL stage.

## Methods

### Participant inclusion

This study was approved by the Mass General Brigham Human Research Committee, and each participant provided written informed consent. From 2016 to 2022, participants with systemic AL amyloidosis were recruited from Brigham and Women’s Hospital (Boston) and from other specialized centres into a prospective cohort study: Molecular Imaging of Primary Amyloid Cardiomyopathy (MICA; NCT02641145).(12–16) Systemic AL amyloidosis was diagnosed by standard criteria, including biopsy with confirmation of amyloid type by immunohistochemistry or mass spectrometry.(17) Key exclusion criteria were significant non-amyloid cardiac disease (e.g., coronary artery disease, severe valvular disease), cardiac pacemaker, implanted cardioverter-defibrillator, estimated glomerular filtration rate (eGFR) <30 mL/min/1.73m^2^ and severe claustrophobia. All participants from the MICA study with recently diagnosed AL amyloidosis at therapy initiation and with baseline MRI were included (N=80), after exclusion of 25 participants in haematological remission and 1 not tolerating MRI. Presence of cardiomyopathy (AL-CMP) was based on sex-specific elevated cardiac troponin T (>0.014 ng/mL in males, or >0.009 ng/mL in females) and/or age-specific elevated N-terminal pro-B-type natriuretic peptide (NT-proBNP, >450 pg/mL if <50 years, >900 pg/mL if 50-75 years, or >1800 pg/mL if >75 years), as recommended and previously done for AL amyloidosis.(14,17) Absence of cardiomyopathy (AL-non-CMP) was defined by troponin T and NT-proBNP under these thresholds, and left ventricular (LV) wall thickness <12 mm in echocardiography or MRI. Participants underwent comprehensive assessments including clinical parameters, cardiac biomarkers and MRI at baseline, 6 months and 12 months for AL-CMP participants (Supplemental Figure 1). AL-non-CMP participants only underwent 6-month follow-up, as no further myocardial change was expected in participants without baseline cardiomyopathy and for whom haematological remission was expected at 6 months. The Mayo AL stage (I-IV) was calculated starting at one point, with one more point assigned for each criterion: troponin T ≥0.025 ng/mL, NT-proBNP ≥1800 pg/mL and difference between involved and uninvolved immunoglobulin free light chains (dFLC) ≥180 mg/L.(18)

### Response to therapy and event ascertainment

Hematologic response was adjudicated by an expert haematologist (GB) as recommended:(19) complete response if dFLC, κ/λ ratio, serum and urine immunofixation were normal, very good partial response if dFLC decreased to <40 mg/mL, partial response if dFLC decreased by >50% to ≥40 mg/mL, and no response if dFLC decreased by ≤50% or did not decrease. ECV response was defined as an absolute decrease or increase by ≥5%, as previously done based on repeatability data.(11,20) Major cardiac events (MACE) were defined as a composite of all-cause death, heart failure hospitalization, or cardiac transplantation. Event ascertainment was performed by phone calls to participants and review of electronic medical records, with 99% successful follow-up.

### Cardiac MRI

Myocardial characteristics, cardiac structure and function were analysed by contrast-enhanced MRI. All exams were performed on a 3 T system (Siemens, Erlangen, Germany) at the Brigham and Women’s Hospital, as previously described.(13,14,16) Our protocol included steady-state free-precession cine imaging for cardiac structure and function (repetition time 3.4 ms; echo time 1.2 ms; temporal resolution 40-50 ms; in-plane spatial resolution 1.5-1.8 x 1.8-2.1 mm) for a standard stack of short-axis slices (thickness 8 mm, no gap) and 3 long-axis planes. A real-time acquisition was used for participants with highly irregular heart rhythm. Myocardial LV native T1 maps were acquired in 3 short-axis slices at the base, mid and apical levels using the modified Look-Locker inversion (MOLLI) recovery technique at end-diastole for 3 cycles (5-3-1 R-R durations with 3-4 R-R rest periods) during a single breath hold. T1 mapping was repeated with the same settings 10 and 20 minutes after intravenous administration of 0.1 mmol/kg of gadolinium contrast agent (gadoterate meglumine, Guerbet LLC, Bloomington, IN, USA, in nearly all patients). T2 mapping was performed before contrast administration using a T2 preparation with varying echo times followed by a single-shot readout of the images in 3 short-axis slices. In-line motion correction was applied to T1 and T2 maps during image reconstruction. A phase-sensitive, inversion recovery-prepared, fast gradient echo sequence, triggered every other heartbeat, was used to assess for LGE in short-axis and long-axis slices 5-10 minutes after contrast injection (appropriate timing for cardiac amyloidosis). Dedicated software was used for postprocessing and measurements (MedisSuite version 3.2, Medical Imaging Systems, Leiden, the Netherlands). The ECV fraction was estimated by plotting the reciprocal of each segmental myocardial T1 against the reciprocal of the blood pool T1 for native, 10-minute and 20-minute post-contrast MOLLI acquisitions, then calculating linear regression slopes, corresponding to segmental partition coefficients for gadolinium (λ_Gd_). Each λ_Gd_ was multiplied by (1 – haematocrit) to obtain segmental ECV fractions, and segmental values were averaged into the global LV ECV fraction, as previously described.(21) This regression-based method over multiple time points reduces measurement variability and offers excellent intra-observer, inter-observer and test-retest absolute agreements for ECV.(22) We evaluated LV myocardial characteristics by ECV, native T1, T2 and typical LGE (diffuse subendocardial or transmural), structural changes by LV mass, right ventricular (RV) mass, left atrial (LA) volume and right atrial (RA) surface, and functional changes by LV stroke volume (SV), LV ejection fraction (EF) and RV EF. Recently updated, sex-specific normal reference values were used.(23) Measurements were performed by experienced readers, showing excellent inter-observer reliability by intraclass correlation coefficients: LV native T1 0.99 (95% confidence interval [CI] 0.97-1.00), LV ECV 0.99 (95%CI 0.97-1.00), LV mass 0.97 (95%CI 0.64-0.99) and LV EF 0.95 (95%CI 0.87-0.98).

### Echocardiography

All participants underwent echocardiography, but only exams performed within 6 months of MRI were included (median absolute time difference 9 days, interquartile range [IQR] 1-48, 12 exams excluded). LV global longitudinal strain (GLS), E/a ratio and E/e’ ratio were measured using dedicated software (TOMTEC Image Arena version 4.6, TOMTEC Imaging Systems GmbH, Unterschleissheim, Germany). We used a normal threshold for LV GLS (-16%) based on recommended values.(24)

### Statistical analysis

Continuous variables were presented as median with IQR and compared across groups using Wilcoxon rank-sum test. Categorical variables were displayed as frequency with percentage and compared using Fisher’s exact test. Correlations were quantified using Spearman’s ρ. Cardiac structural metrics were indexed by body surface area for comparisons between groups at baseline, but not for analyses over time, because changes in body weight would affect indexed values even without structural changes. Temporal changes were analysed over visits using Wilcoxon signed-rank test (paired), and further analysed over time using linear mixed-effects regression with random intercepts by participant, random slopes for time and the Kenward-Roger method. For the outcome analysis, Cox regression was performed after checking the related assumptions. Prediction performance of MRI metrics was compared using Akaike’s information criterion (AIC) and the likelihood-ratio test for nested models on participants without missing values.(25) Kaplan-Meier analysis was conducted with the log-rank test, and presented for the MRI metric with the best outcome prediction. Level cut-offs were based on normal reference values and on log-rank statistic maximization, with a supplemental analysis based on tertiles. Event rates, accounting for multiple events in the same participant, were compared using an exact test with Poisson distribution. We presented two-sided p-values and considered them statistically significant if <0.05. Adjustment for multiple testing was performed with the Benjamini-Hochberg procedure. Data were analysed using R version 4.3.1 (R Core Team. R: A language and environment for statistical computing. R Foundation for Statistical Computing, Vienna, Austria) and the packages *tidyverse*, *DescTools*, *gtsummary*, *rstatix*, *correlation*, *lme4*, *pbkrtest*, *survival*, *survminer* and *ggsurvplot*. Results were reported according to the STROBE statement.(26)

## Results

### Baseline characteristics

We included 80 participants: 60 AL-CMP (75%) and 20 AL-non-CMP (25%). There were 46 males (58%) and the median age was 62 years (IQR 57-67). Compared with AL-non-CMP, AL-CMP participants were older, had lower blood pressure, worse laboratory values, Mayo AL stage and function on echocardiography at baseline, and received different therapies for AL amyloidosis (Table 1).

**Table 1.** Participant Characteristics at Baseline.

| Variable | AL-CMP<br>(N = 60) | AL-non-CMP<br>(N = 20) | p-value |
| --- | --- | --- | --- |
| **Demographic variables** |  |  |  |
| Age (years) | 63 (59 – 68) | 59 (51 – 65) | 0.046 |
| Male sex | 32 (53%) | 14 (70%) | 0.296 |
| Race and ethnicity |  |  | 0.339 |
| White Non-Hispanic | 52 (87%) | 16 (80%) |  |
| Black or African American | 5 (8.3%) | 4 (20%) |  |
| Other | 3 (5.0%) | 0 (0%) |  |
| **Clinical parameters** |  |  |  |
| Body mass index (kg/m<sup>2</sup>) | 25.4 (23.0 – 27.6) | 24.7 (23.3 – 28.5) | 0.726 |
| Heart rate (bpm) | 80 (74 – 86) | 75 (68 – 83) | 0.203 |
| Systolic blood pressure (mmHg) | 107 (97 – 120) | 127 (106 – 135) | 0.003 |
| Diastolic blood pressure (mmHg) | 66 (60 – 72) | 68 (62 – 73) | 0.383 |
| **Laboratory tests** |  |  |  |
| Haemoglobin (g/dL) | 13.1 (11.4 – 13.9) | 13.5 (13.1 – 14.5) | 0.064 |
| eGFR (mL/min/1.73m<sup>2</sup>) | 68 (52 – 80) | 89 (74 – 100) | 0.002 |
| Troponin T (ng/mL) | 0.074 (0.033 – 0.109) | 0.009 (0.007 – 0.011) | <0.001 |
| NT-proBNP (pg/mL) | 4420 (1627 – 8451) | 122 (64 – 268) | <0.001 |
| dFLC (mg/L) | 276 (101 – 502) | 122 (71 – 264) | 0.044 |
| Mayo AL stage | 3 (3 – 4) | 1 (1 – 2) | <0.001 |
| **Echocardiography** |  |  |  |
| LV GLS (%) | -13.8 (-17.2 – -11.5) | -23.3 (-25.1 – -20.5) | <0.001 |
| Abnormal LV GLS (>-16%) | 39 (68%) | 0 (0%) | <0.001 |
| E/A ratio | 2.26 (1.50 – 3.27) | 0.93 (0.86 – 1.06) | <0.001 |
| E/e' ratio | 20.9 (16.6 – 25.9) | 7.1 (6.0 – 10.3) | <0.001 |
| **Treatment at and after baseline** |  |  |  |
| CyBorD | 50 (83%) | 8 (40%) | <0.001 |
| Daratumumab | 31 (52%) | 10 (50%) | >0.999 |
| Other chemotherapy | 21 (35%) | 13 (65%) | 0.035 |
| ASCT | 6 (10%) | 11 (55%) | <0.001 |

Continuous variables were presented as median (interquartile range) and compared using Wilcoxon rank-sum test. Categorical variables were presented as frequencies (percentages) and compared using Fisher's exact test.
AL-CMP/AL-non-CMP: Light-chain amyloidosis with/without cardiomyopathy. ASCT: Autologous stem cell transplantation. CyBorD: Cyclophosphamide-bortezomib-dexamethasone. dFLC: Difference in immunoglobulin free light chains. eGFR: Estimated glomerular filtration rate. GLS: Global longitudinal strain. LGE: Late gadolinium enhancement. LV: Left ventricular. NT-proBNP: N-terminal pro-B-type natriuretic peptide.

### LV myocardial characteristics at baseline in AL-CMP and AL-non-CMP

As expected, all myocardial characteristics were worse in AL-CMP than in AL-non-CMP at baseline, as well as LV myocardial structure and function (Table 2, Figure 1). In the AL-CMP cohort, median LV ECV was 53% (IQR 47-58), abnormal LV ECV (>32%) and typical LV LGE were found in all participants, while LV GLS on echocardiography was abnormal in 68%. Notably, our study showed abnormal LV myocardial characteristics in nearly half of the AL-non-CMP cohort. Their median LV ECV was 30% (IQR 27-38), LV ECV was abnormal (>32%) in 47%, typical LV LGE was found in 21%, while LV GLS on echocardiography was normal in all participants. These results suggest that myocardial characteristics detect early interstitial changes in AL-non-CMP, prior to overt changes in cardiac structure, cardiac function, or biomarker release. Moreover, we found moderate to strong correlations between LV ECV and multiple cardiac structural and functional metrics (Supplemental Table 1, Supplemental Figure 2).

**Figure 1.**
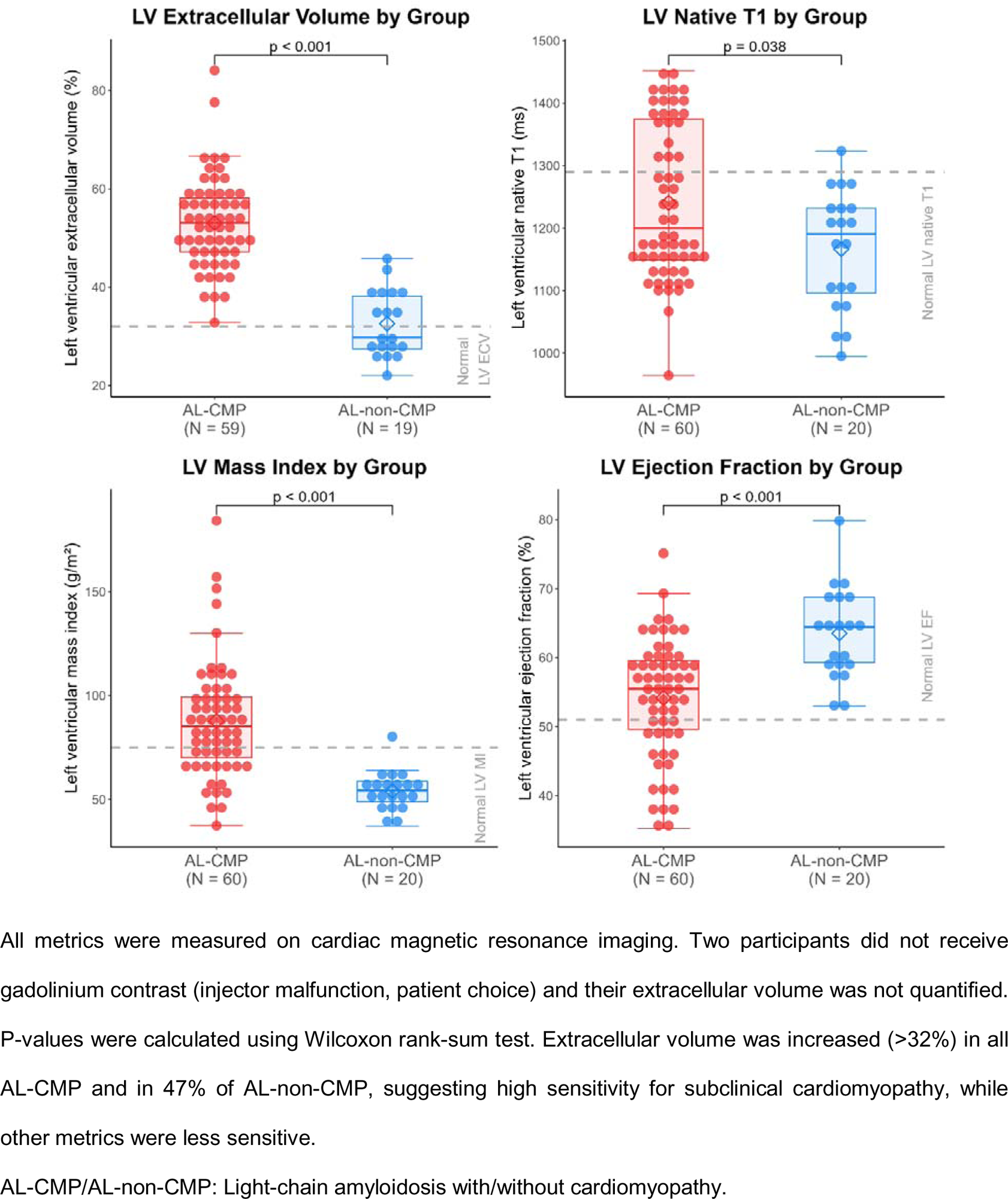
Baseline Myocardial Characteristics, Cardiac Structure and Function.

**Table 2.**
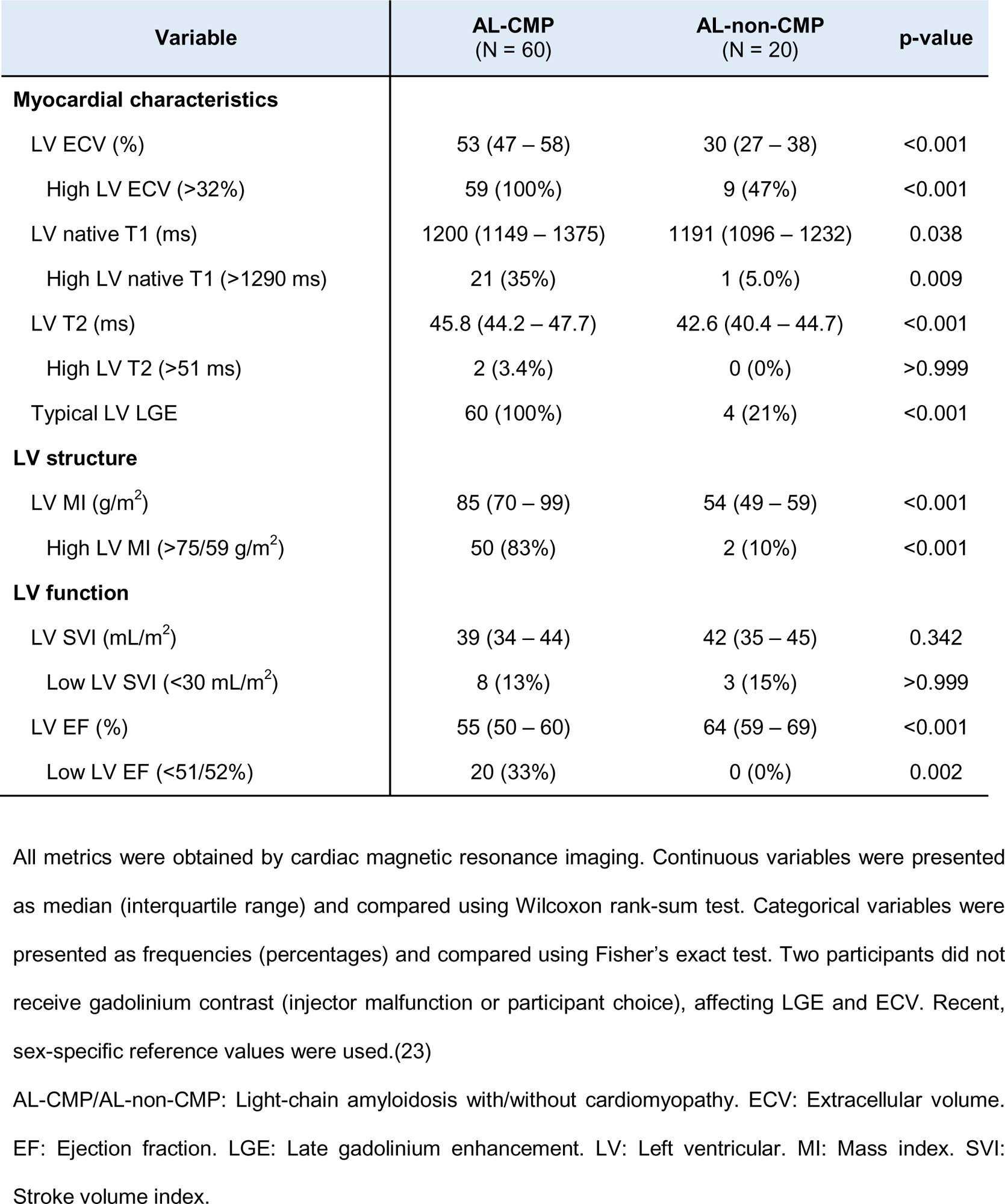
Left Ventricular Myocardial Characteristics, Structure and Function at Baseline.

### Follow-up visits and response data

Fifty-one participants completed the 6-month visit (33 AL-CMP, 18 AL-non-CMP), and 29 completed the 12-month visit (all AL-CMP). Reasons for missed follow-up visits included death, worsening health, COVID-related shutdown, cardiac device implantation, or personal choice (Supplemental Figure 1). Of the returning participants, 87% reached CR or VGPR by 6 months and 93% by 12 months (Table 3).

**Table 3.**
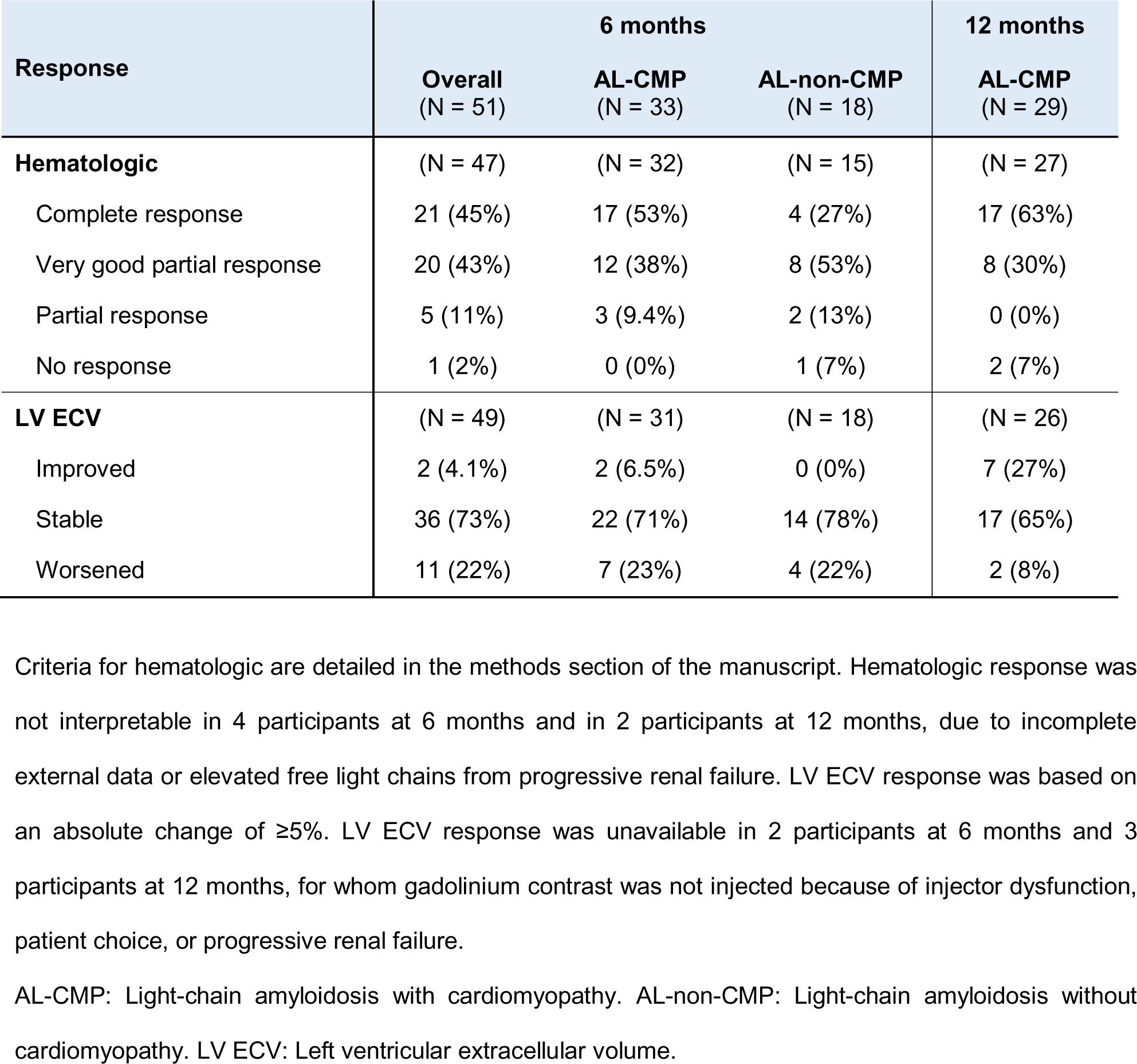
Response Analysis at Follow-up Visits.

| Response | 6 months |  |  | 12 months |
| --- | --- | --- | --- | --- |
|  | Overall<br>(N = 51) | AL-CMP<br>(N = 33) | AL-non-CMP<br>(N = 18) | AL-CMP<br>(N = 29) |
| **Hematologic** | (N = 47) | (N = 32) | (N = 15) | (N = 27) |
| Complete response | 21 (45%) | 17 (53%) | 4 (27%) | 17 (63%) |
| Very good partial response | 20 (43%) | 12 (38%) | 8 (53%) | 8 (30%) |
| Partial response | 5 (11%) | 3 (9.4%) | 2 (13%) | 0 (0%) |
| No response | 1 (2%) | 0 (0%) | 1 (7%) | 2 (7%) |
| **LV ECV** | (N = 49) | (N = 31) | (N = 18) | (N = 26) |
| Improved | 2 (4.1%) | 2 (6.5%) | 0 (0%) | 7 (27%) |
| Stable | 36 (73%) | 22 (71%) | 14 (78%) | 17 (65%) |
| Worsened | 11 (22%) | 7 (23%) | 4 (22%) | 2 (8%) |

Criteria for hematologic are detailed in the methods section of the manuscript. Hematologic response was not interpretable in 4 participants at 6 months and in 2 participants at 12 months, due to incomplete external data or elevated free light chains from progressive renal failure. LV ECV response was based on an absolute change of $\geq 5\%$ . LV ECV response was unavailable in 2 participants at 6 months and 3 participants at 12 months, for whom gadolinium contrast was not injected because of injector dysfunction, patient choice, or progressive renal failure.
AL-CMP: Light-chain amyloidosis with cardiomyopathy. AL-non-CMP: Light-chain amyloidosis without cardiomyopathy. LV ECV: Left ventricular extracellular volume.

### Temporal changes in LV myocardial characteristics

Cardiac MRI was repeated at 6 months and, in AL-CMP, at 12 months. LV MRI structural and functional metrics (LV mass, LV stroke volume index and LV EF) did not change over time, but LV myocardial characteristics showed dynamic temporal changes (Table 4, Figure 2, Figure 3). ECV tended to increase at 6 months in AL-CMP (median +2%, IQR-1 to +5, p=0.120) and AL-non-CMP (+2%, IQR -1 to +3, p=0.018). In AL-CMP, LV ECV decreased from 6 months to 12 months in AL-CMP (-3%, IQR -6 to +1, p=0.011). Native T1 increased at 6 months in AL-CMP (+50 ms, IQR +9 to +113, p=0.003) and AL-non-CMP (+38 ms, IQR +6 to +87, p=0.003), then remained stable at 12 months in AL-CMP (-11 ms, IQR -39 to +31, p=0.853) vs. 6 months. Longitudinal mixed-effects models confirmed the curved trend of LV ECV upwards at 6 months and downwards at 12 months (time p=0.005; time^2^ p<0.001), and confirmed the increase of LV native T1 over time (p=0.001; Figure 3). For most participants, ECV did not substantially change during the study, but 22% exhibited an unfavourable ECV response (+≥5%) at 6 months, and 27% a favourable ECV response (-≥5%) at 12 months, in line with the curved trend (Table 3, Figure 3).

**Figure 2.**
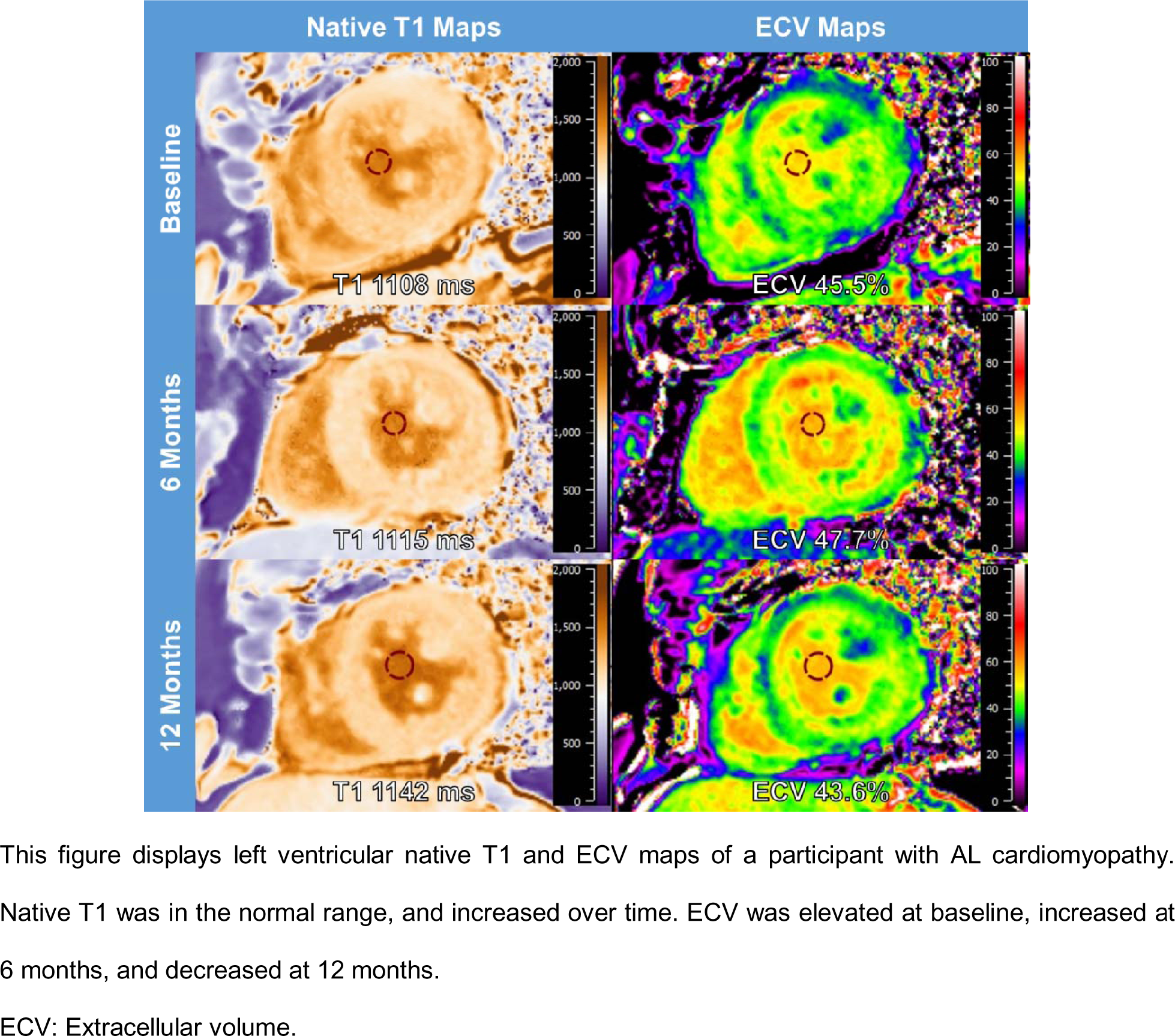
Changes in Myocardial Characteristics over Time.

**Figure 3.**
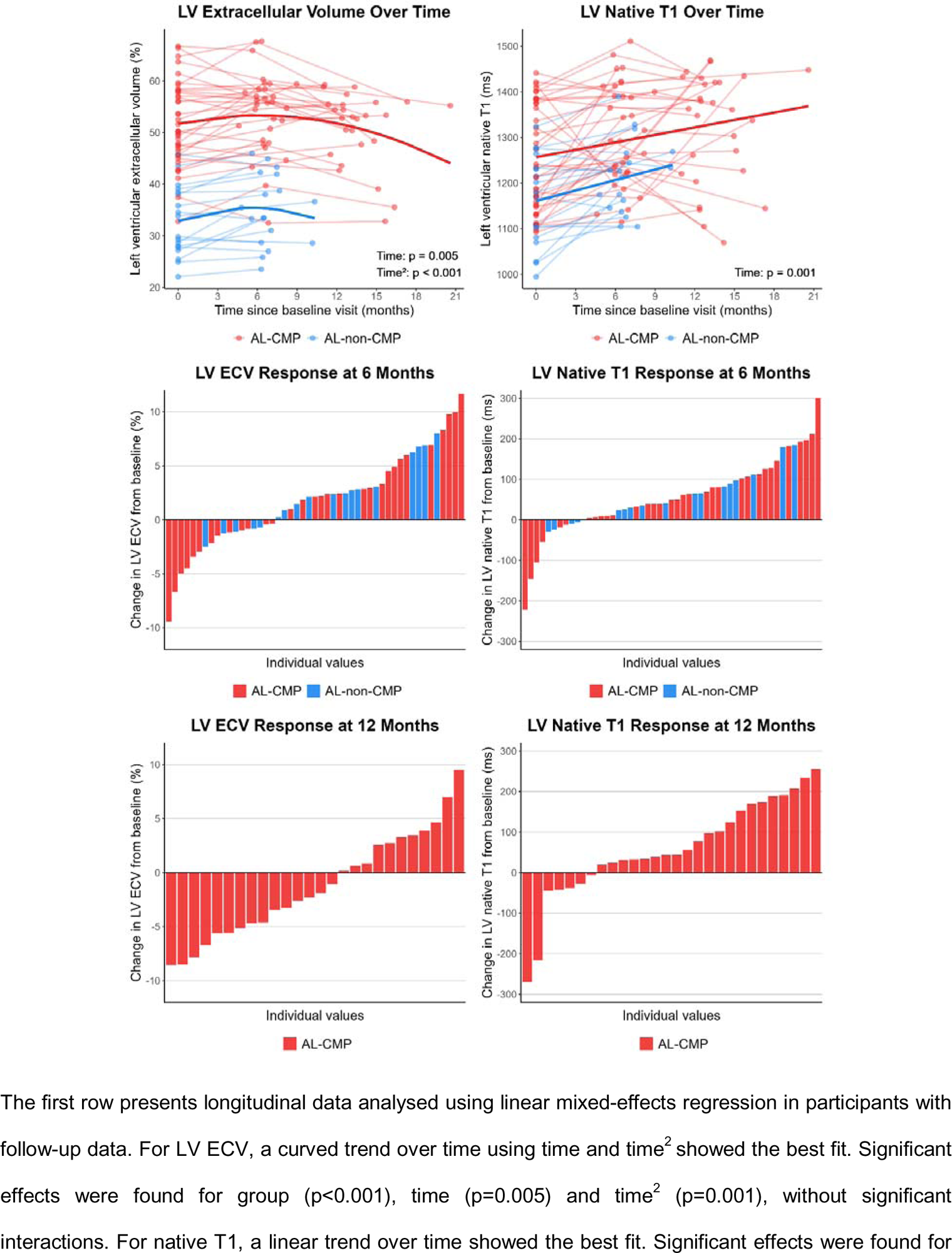
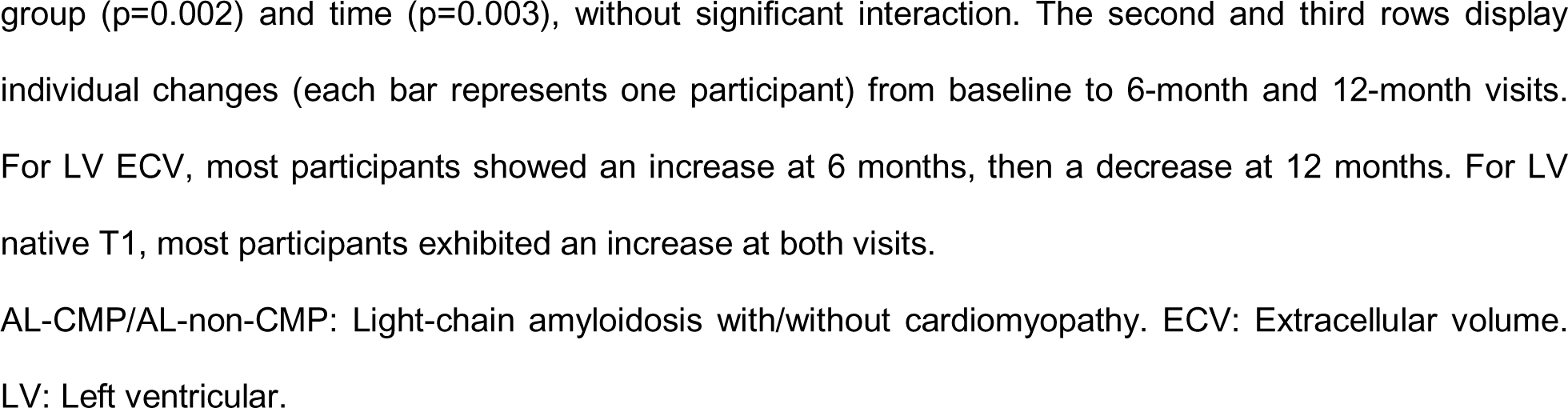
Longitudinal Analysis of Myocardial Characteristics.

**Table 4.**
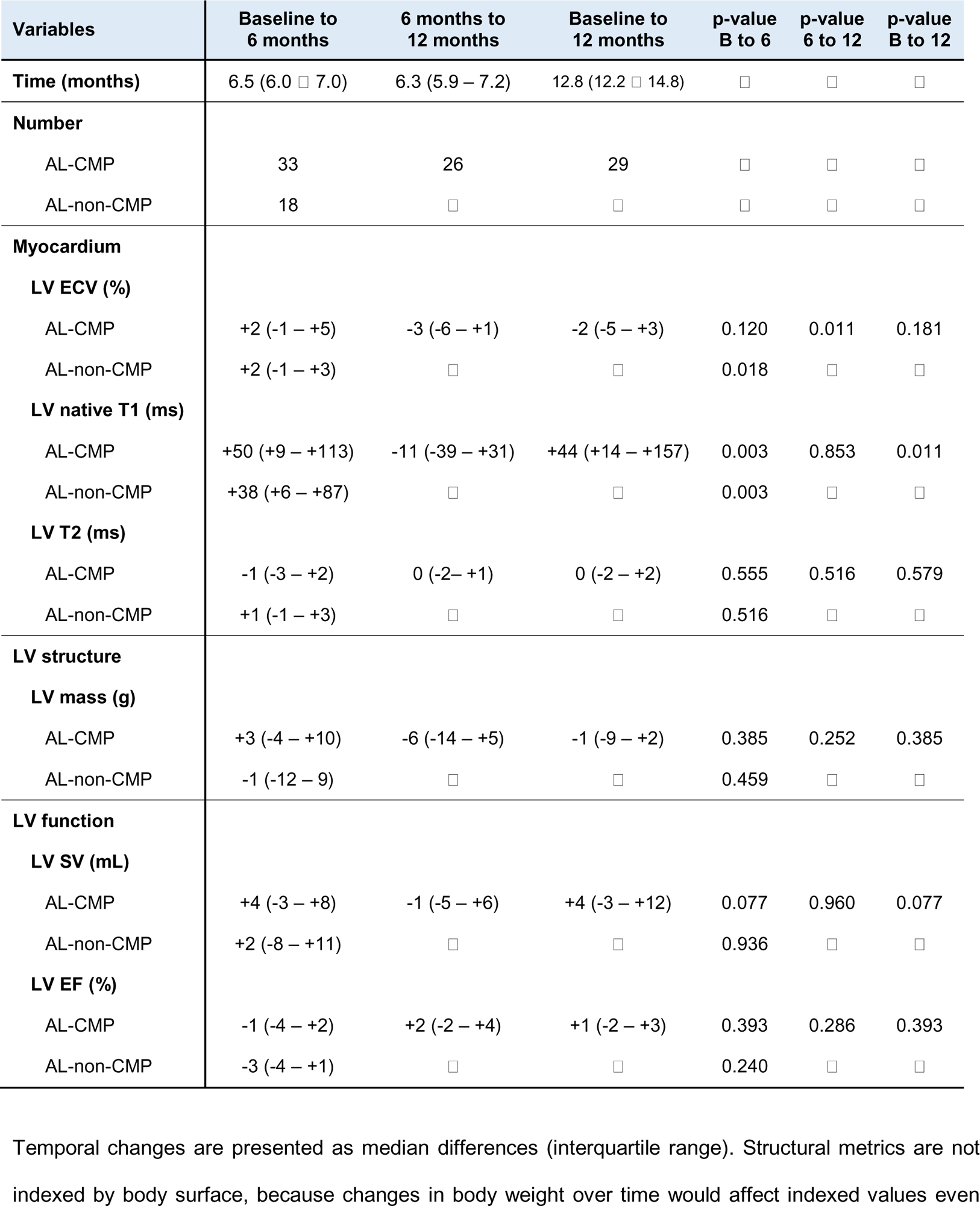
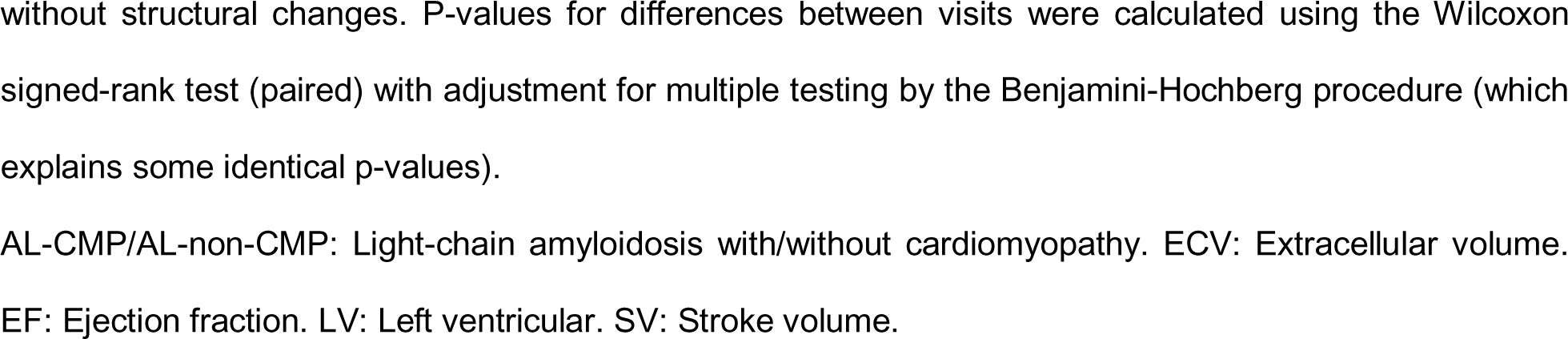
Temporal Changes in Left Ventricular Myocardial Characteristics, Structure and Function Between Visits.

| Variables | Baseline to 6 months | 6 months to 12 months | Baseline to 12 months | p-value B to 6 | p-value 6 to 12 | p-value B to 12 |
| --- | --- | --- | --- | --- | --- | --- |
| **Time (months)** | 6.5 (6.0 – 7.0) | 6.3 (5.9 – 7.2) | 12.8 (12.2 – 14.8) | □ | □ | □ |
| **Number** |  |  |  |  |  |  |
| AL-CMP | 33 | 26 | 29 | □ | □ | □ |
| AL-non-CMP | 18 | □ | □ | □ | □ | □ |
| **Myocardium** |  |  |  |  |  |  |
| **LV ECV (%)** |  |  |  |  |  |  |
| AL-CMP | +2 (-1 – +5) | -3 (-6 – +1) | -2 (-5 – +3) | 0.120 | 0.011 | 0.181 |
| AL-non-CMP | +2 (-1 – +3) | □ | □ | 0.018 | □ | □ |
| **LV native T1 (ms)** |  |  |  |  |  |  |
| AL-CMP | +50 (+9 – +113) | -11 (-39 – +31) | +44 (+14 – +157) | 0.003 | 0.853 | 0.011 |
| AL-non-CMP | +38 (+6 – +87) | □ | □ | 0.003 | □ | □ |
| **LV T2 (ms)** |  |  |  |  |  |  |
| AL-CMP | -1 (-3 – +2) | 0 (-2 – +1) | 0 (-2 – +2) | 0.555 | 0.516 | 0.579 |
| AL-non-CMP | +1 (-1 – +3) | □ | □ | 0.516 | □ | □ |
| **LV structure** |  |  |  |  |  |  |
| **LV mass (g)** |  |  |  |  |  |  |
| AL-CMP | +3 (-4 – +10) | -6 (-14 – +5) | -1 (-9 – +2) | 0.385 | 0.252 | 0.385 |
| AL-non-CMP | -1 (-12 – 9) | □ | □ | 0.459 | □ | □ |
| **LV function** |  |  |  |  |  |  |
| **LV SV (mL)** |  |  |  |  |  |  |
| AL-CMP | +4 (-3 – +8) | -1 (-5 – +6) | +4 (-3 – +12) | 0.077 | 0.960 | 0.077 |
| AL-non-CMP | +2 (-8 – +11) | □ | □ | 0.936 | □ | □ |
| **LV EF (%)** |  |  |  |  |  |  |
| AL-CMP | -1 (-4 – +2) | +2 (-2 – +4) | +1 (-2 – +3) | 0.393 | 0.286 | 0.393 |
| AL-non-CMP | -3 (-4 – +1) | □ | □ | 0.240 | □ | □ |

Temporal changes are presented as median differences (interquartile range). Structural metrics are not indexed by body surface, because changes in body weight over time would affect indexed values even
without structural changes. P-values for differences between visits were calculated using the Wilcoxon signed-rank test (paired) with adjustment for multiple testing by the Benjamini-Hochberg procedure (which explains some identical p-values).
AL-CMP/AL-non-CMP: Light-chain amyloidosis with/without cardiomyopathy. ECV: Extracellular volume.
EF: Ejection fraction. LV: Left ventricular. SV: Stroke volume.

### Prediction of adverse outcomes by LV myocardial characteristics

During a median follow-up of 27 months (IQR 3-45), 39 participants experienced MACE (49%), among which 22 died (28%), 24 were hospitalized for heart failure (30%), and 3 underwent cardiac transplantation (3.8%). Notably, 24 MACE (62%) occurred in the first 6 months. All events were substantially more frequent in AL-CMP than in AL-non-CMP (Supplemental Table 2). In univariable analysis, LV ECV and typical LGE associated with MACE and death, but native T1 and T2 were not (Table 5). Based on AIC (lower is better), LV ECV was the best predictor of MACE and death (AIC 270 and 149, both p<0.001). In Kaplan-Meier analysis and event rate analysis, LV ECV levels were excellent predictors of MACE and death (both p<0.001 for trend; Figure 4). Normal LV ECV ≤32% identified a cohort without any MACE during the follow-up, while high LV ECV >48% (by log-rank maximization) identified a high-risk cohort with 74% MACE. Among AL-non-CMP, all MACE occurred in participants with abnormal LV ECV, without typical LGE. For death, the optimal LV ECV cut-off was >60%, identifying a cohort with 82% death. Similar results were found using LV ECV tertiles (Supplemental Figure 3). In multivariable analysis combining each MRI metric with the Mayo AL stage, only LV ECV remained significantly associated with both MACE and death (Table 5). LV ECV offered the best improvement in prediction for MACE and death over the Mayo AL stage alone (AIC 269 and 147, both likelihood-ratio p≤0.01).

**Figure 4.**
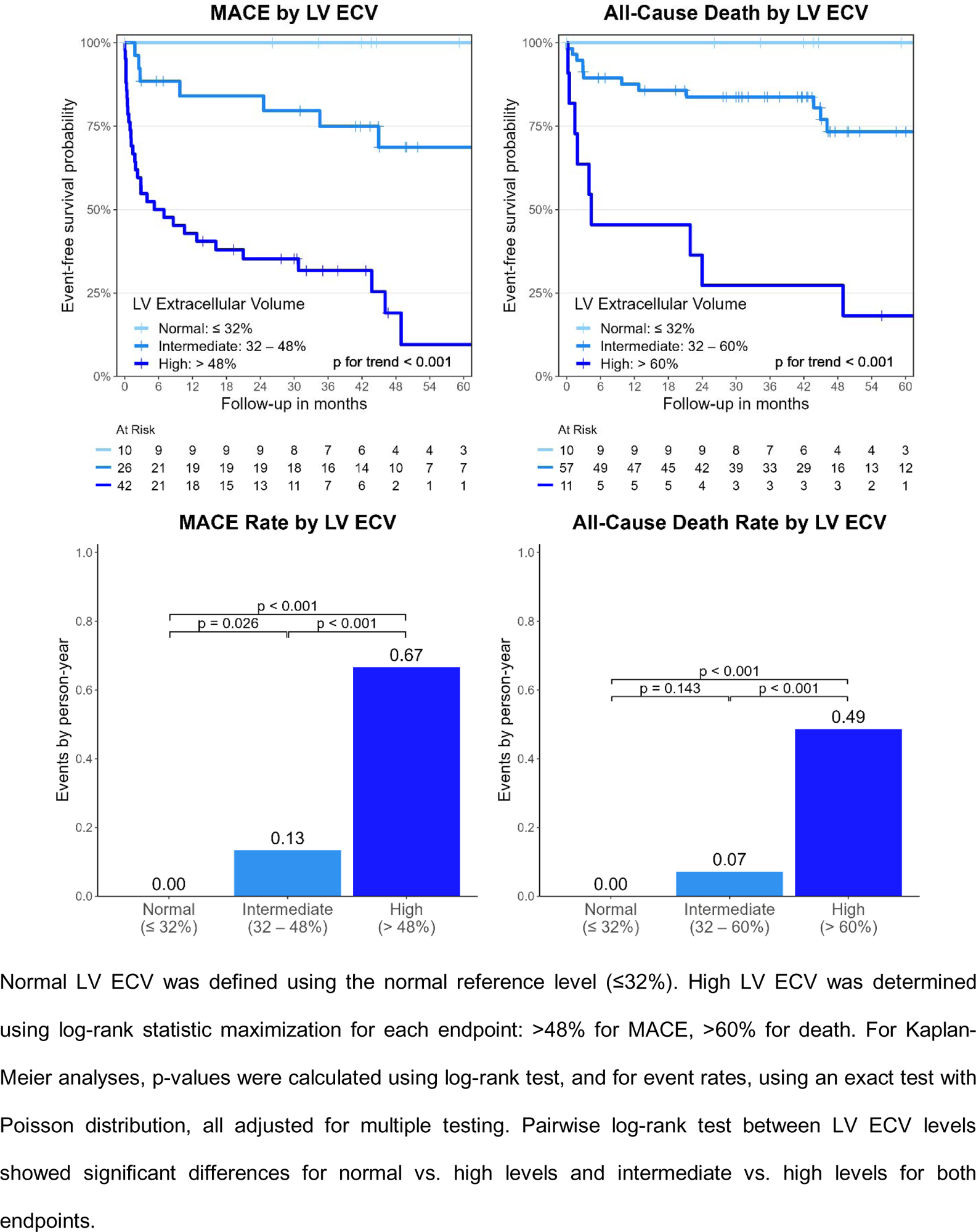
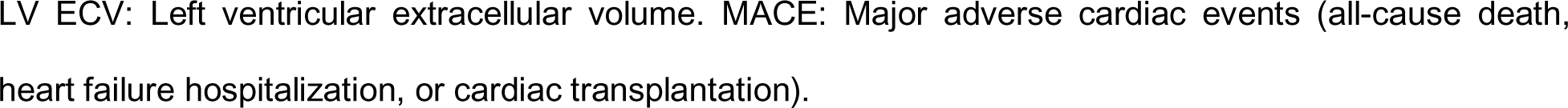
Survival Analysis Based on Left Ventricular Interstitial Expansion.

**Table 5.**
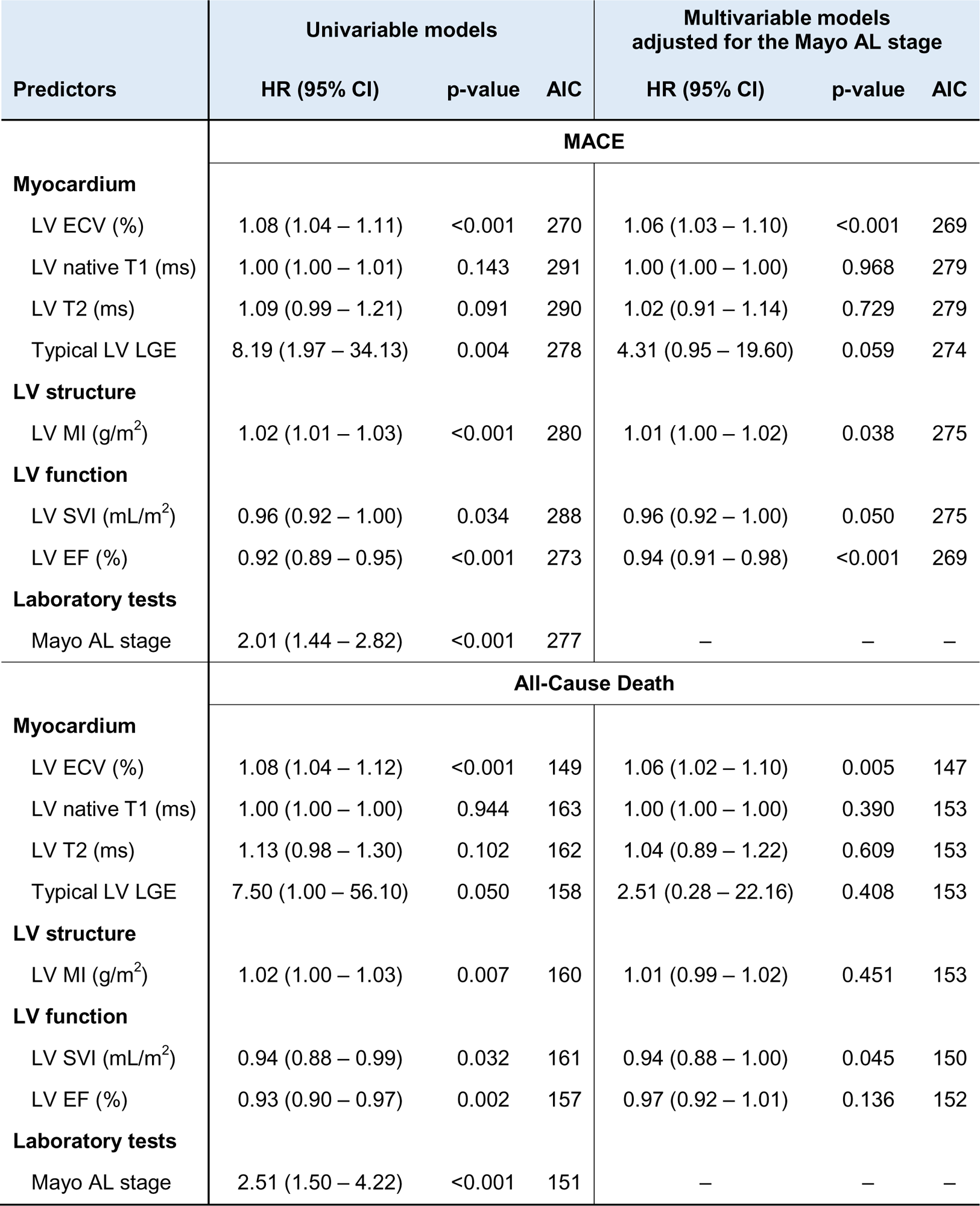
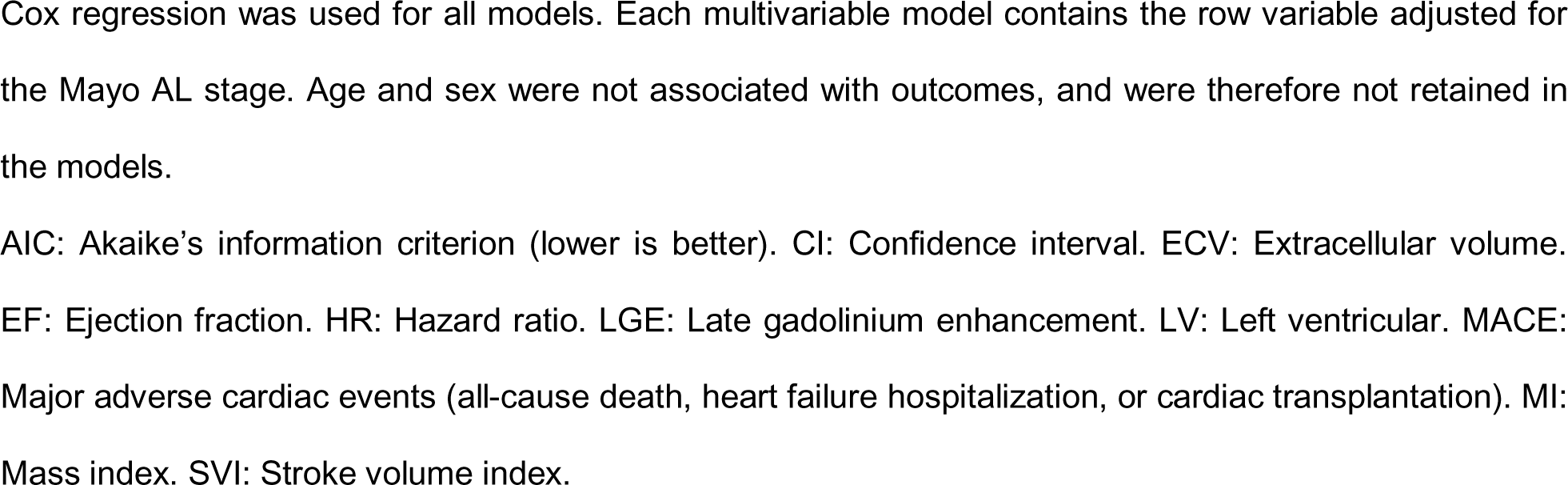
Association of Left Ventricular Myocardial Characteristics with MACE and All-Cause Death.

| Predictors | Univariable models |  |  | Multivariable models<br>adjusted for the Mayo AL stage |  |  |
| --- | --- | --- | --- | --- | --- | --- |
|  | HR (95% CI) | p-value | AIC | HR (95% CI) | p-value | AIC |
| **MACE** |  |  |  |  |  |  |
| **Myocardium** |  |  |  |  |  |  |
| LV ECV (%) | 1.08 (1.04 – 1.11) | <0.001 | 270 | 1.06 (1.03 – 1.10) | <0.001 | 269 |
| LV native T1 (ms) | 1.00 (1.00 – 1.01) | 0.143 | 291 | 1.00 (1.00 – 1.00) | 0.968 | 279 |
| LV T2 (ms) | 1.09 (0.99 – 1.21) | 0.091 | 290 | 1.02 (0.91 – 1.14) | 0.729 | 279 |
| Typical LV LGE | 8.19 (1.97 – 34.13) | 0.004 | 278 | 4.31 (0.95 – 19.60) | 0.059 | 274 |
| **LV structure** |  |  |  |  |  |  |
| LV MI (g/m<sup>2</sup>) | 1.02 (1.01 – 1.03) | <0.001 | 280 | 1.01 (1.00 – 1.02) | 0.038 | 275 |
| **LV function** |  |  |  |  |  |  |
| LV SVI (mL/m<sup>2</sup>) | 0.96 (0.92 – 1.00) | 0.034 | 288 | 0.96 (0.92 – 1.00) | 0.050 | 275 |
| LV EF (%) | 0.92 (0.89 – 0.95) | <0.001 | 273 | 0.94 (0.91 – 0.98) | <0.001 | 269 |
| **Laboratory tests** |  |  |  |  |  |  |
| Mayo AL stage | 2.01 (1.44 – 2.82) | <0.001 | 277 | – | – | – |
| **All-Cause Death** |  |  |  |  |  |  |
| **Myocardium** |  |  |  |  |  |  |
| LV ECV (%) | 1.08 (1.04 – 1.12) | <0.001 | 149 | 1.06 (1.02 – 1.10) | 0.005 | 147 |
| LV native T1 (ms) | 1.00 (1.00 – 1.00) | 0.944 | 163 | 1.00 (1.00 – 1.00) | 0.390 | 153 |
| LV T2 (ms) | 1.13 (0.98 – 1.30) | 0.102 | 162 | 1.04 (0.89 – 1.22) | 0.609 | 153 |
| Typical LV LGE | 7.50 (1.00 – 56.10) | 0.050 | 158 | 2.51 (0.28 – 22.16) | 0.408 | 153 |
| **LV structure** |  |  |  |  |  |  |
| LV MI (g/m<sup>2</sup>) | 1.02 (1.00 – 1.03) | 0.007 | 160 | 1.01 (0.99 – 1.02) | 0.451 | 153 |
| **LV function** |  |  |  |  |  |  |
| LV SVI (mL/m<sup>2</sup>) | 0.94 (0.88 – 0.99) | 0.032 | 161 | 0.94 (0.88 – 1.00) | 0.045 | 150 |
| LV EF (%) | 0.93 (0.90 – 0.97) | 0.002 | 157 | 0.97 (0.92 – 1.01) | 0.136 | 152 |
| **Laboratory tests** |  |  |  |  |  |  |
| Mayo AL stage | 2.51 (1.50 – 4.22) | <0.001 | 151 | – | – | – |

Cox regression was used for all models. Each multivariable model contains the row variable adjusted for the Mayo AL stage. Age and sex were not associated with outcomes, and were therefore not retained in the models.
AIC: Akaike's information criterion (lower is better). CI: Confidence interval. ECV: Extracellular volume. EF: Ejection fraction. HR: Hazard ratio. LGE: Late gadolinium enhancement. LV: Left ventricular. MACE: Major adverse cardiac events (all-cause death, heart failure hospitalization, or cardiac transplantation). MI: Mass index. SVI: Stroke volume index.

## Discussion

This study revealed several novel and intriguing findings on myocardial characteristics by MRI in systemic AL amyloidosis. First, at baseline, myocardial characteristics were worse in AL-CMP than in AL-non-CMP, as expected. But among AL-non-CMP participants, 47% had abnormal LV ECV, despite normal cardiac biomarkers and LV wall thickness recommended to define cardiac involvement in AL amyloidosis.(17) Moreover, GLS was abnormal in no participant of the AL-non-CMP cohort, supporting that abnormal ECV is an early myocardial interstitial change which has not yet impacted myocardial contractile function or provoked cardiac biomarker release, which are markers of poor prognosis. This suggests a superior sensitivity of LV ECV to identify early cardiac involvement in AL amyloidosis. Second, the longitudinal analysis after plasma cell therapy initiation demonstrated a curved evolution of LV ECV over time, with an increasing trend at 6 months, and a decrease from 6 to 12 months, highlighting temporal variations in interstitial expansion. Moreover, LV native T1 increased at 6 months, then remained stable at 12 months, reflecting remodelling of the interstitial and/or intracellular compartments. However, T2 did not change over time, suggesting no relevant contribution of oedema. These alterations paralleled a favourable haematological response at 6 and 12 months. Third, in outcome analysis, LV ECV emerged as the best predictor of MACE and death. LV ECV showed an excellent risk stratification capacity, identifying cohorts of participants at very low or very high risk of adverse outcomes. In multivariable analysis, LV ECV offered additional prognostic performance when combined with the Mayo AL stage.

An important novel finding of this study, which was not clearly addressed in prior works, is the prospective evaluation of myocardial characteristics in AL amyloidosis participants with and without cardiomyopathy (AL-CMP, AL-non-CMP). For AL-CMP, our findings of myocardial alterations are concordant with previous data.(9,11) But in our study, cardiac MRI also identified abnormal LV ECV in about half of participants diagnosed as AL-non-CMP by conventional criteria (normal wall thickness, troponin T and NT-proBNP).(17) Other imaging metrics of cardiac amyloid infiltration, such as LV LGE, native T1, or GLS on echocardiography did not identify as many abnormal cases among AL-non-CMP, suggesting a higher sensitivity of LV ECV for subclinical cardiomyopathy in AL amyloidosis, using current reference values.(23) A previous study from the National Amyloidosis Centre (NAC) in the United Kingdom found a lower proportion of abnormal LV ECV in AL-non-CMP, but a 1.5 T MRI scanner was used, a different method was utilized to quantify ECV, and ECV was characterized only in the basal interventricular septum.(9) However, the presence of amyloid deposits in about half of AL-non-CMP is consistent with recent findings based on the amyloid-specific radiotracer ^18^F-florbetapir in positron emission tomography.(14) Interestingly, in the present study, all AL-non-CMP participants who experienced MACE had elevated LV ECV, supporting the clinical relevance of this finding. Therefore, the biomarker-based criteria for cardiomyopathy in AL amyloidosis should not be taken alone, but rather complemented with imaging-based criteria, particularly LV ECV. Our findings imply that cardiac MRI with LV ECV estimation could be considered in all newly diagnosed patients with systemic AL amyloidosis.

Temporal changes in myocardial characteristics with plasma cell therapy for systemic AL amyloidosis have not been thoroughly assessed. Our findings of LV ECV following a curved trend upwards at 6 months and downwards at 12 months are consistent with recent results from the NAC cohort.(11) Our study strengthens this finding using a prospective and structured cohort design, additionally including AL-non-CMP. The curved trend of LV interstitial expansion may be due to an initially continued amyloid deposition in the myocardium, followed by amyloid clearance, as suggested by lower ECV at 2 years in the NAC cohort.(11) However, ECV expansion is not specific for amyloid, and this initial increase, more significant in AL-non-CMP, may reflect fibrosis, inflammation, apoptosis or other forms of adverse myocardial remodeling.(27) In the present study, a significant increase of LV native T1 was observed in AL-CMP and AL-non-CMP, but in the NAC study, such an increase was only significant in participants with ECV progression at 12 months.(11) These different changes in ECV and native T1 have important implications for understanding alterations in myocardial composition associated with haematological remission, as they imply a complex remodelling of the myocardial contents. While ECV reflects primarily a structural characteristic of the myocardium, native T1 provides additional information about changes in the interstitial and intracellular composition. An increase of native T1 is usually observed with an expansion of the extracellular space, as the extracellular T1 is generally longer than intracellular T1. This T1 increase from interstitial expansion predominates in cardiac amyloidosis, but the accumulation of amyloid fibrils may ultimately decrease interstitial T1.(28) This effect of amyloid fibrils may explain why native T1 stabilizes while ECV starts to decrease, if this is due to a reduction in amyloid burden. Further alterations in extracellular and intracellular composition may also contribute to changes in native T1. Moreover, persistent interstitial expansion at 12 months, as shown by abnormal LV ECV at 12 months in all participants with AL-CMP, and high early mortality despite state-of-the-art effective therapies,(3) highlight the need for novel, amyloid-reducing treatments in AL amyloidosis.(29,30) For such therapies, measurements of myocardial characteristics may identify eligible patients, measure treatment efficacy, and serve as surrogate outcome measures.

In systemic AL amyloidosis, cardiac involvement portends poor prognosis. Myocardial characteristics from cardiac MRI, particularly LV ECV, have been associated with death.(4,5,7,8) But their association with MACE and their ability to improve prediction beyond the Mayo AL stage had not been clearly established.(7,8) Our study adds new evidence on these topics, showing that LV ECV predicts both MACE and death. LV ECV offered superior prognostic performance for both endpoints, even when combined with the Mayo AL stage. Moreover, the ability of LV ECV ≤32% to identify low-risk patients, while values >48% identify high risk of MACE, and >60% high risk of death, may henceforth contribute to personalized decision-making in the treatment of AL amyloidosis. Unlike previous works,(5,6,8) we found no association between LV native T1 or T2 and outcomes. This may be due to our inclusion of participants with newly diagnosed AL amyloidosis only, or to the use of 3 T MRI. Notably, although LV EF provided independent prognostic value when combined with the Mayo AL stage, it did not identify early cardiac AL amyloidosis and did not change in response to therapy, highlighting the key role for myocardial characterization with ECV in this disease. These results suggest that LV ECV could become part of an updated prognostic staging system.

Our study has some limitations. First, the small sample size for follow-up visits limited our statistical power to analyze changes in cardiac MRI metrics. However, our statistically significant results support large effect sizes with clinical relevance. Our cohort is also among the largest on systemic AL amyloidosis, which is a rare disease. Second, selection bias through survival affected our longitudinal analyses, as a substantial fraction of participants missed follow-up visits, mostly due to death or illness. Thus, changes over time and response data were based on participants with better disease outcomes, who were able to attend follow-up visits. This limitation is unavoidable in a longitudinal study of a disease with high morbidity and mortality. Because data missingness was not at random, we did not use data imputation. Third, participants remaining in the study without reaching CR or VGPR were too few to analyse the effect of hematologic remission on longitudinal analyses or outcomes. This highlights the efficacy of current therapy. However, our study also has important strengths, such as the inclusion of participants with AL amyloidosis at therapy initiation, including AL-non-CMP participants, the prospective, structured cohort design, with extensive assessments including clinical data, biomarkers and imaging, and the comparatively long follow-up duration.

In conclusion, myocardial interstitial expansion estimated by LV ECV is a sensitive marker to detect subclinical cardiac involvement, a quantitative marker to monitor changes in response to plasma cell therapy, and a powerful marker to assess prognosis in addition to the Mayo AL stage in patients with systemic AL amyloidosis.

## Data Availability

All data produced in the present study are available upon reasonable request to the authors.

## Acknowledgments

We are extremely grateful to each of the study participants and their families for their participation and to our funding partners for making this study possible.

## Funding

This work was supported by the National Institutes of Health.

Dorbala: R01 HL 130563; K24 HL 157648; AHA16 CSA 2888 0004; AHA19SRG34950011

Falk: R01 HL 130563

Liao: AHA16 CSA 2888 0004; AHA19SRG34950011

Ruberg: R01 HL 130563; R01 HL 093148

https://clinicaltrials.gov/ct2/show/NCT02641145

## Conflicts of Interest

Clerc: Research fellowship from the International Society of Amyloidosis and Pfizer. Cuddy: Investigator-initiated research grant from Pfizer.

Ruberg: Consulting fees from Astra Zeneca, research support from Pfizer, Alnylam, and Ionis/Akcea.

DiCarli: Research grant from Spectrum Dynamics and Gilead, consulting fees from Sanofi and General

Electric.

Kwong: Grant funding from Alynlam Pharmaceuticals.

Falk: Consulting fees from Ionis Pharmaceuticals, Alnylam Pharmaceuticals, Caelum Biosciences, research funding from GlaxoSmithKline and Akcea.

Dorbala: Consulting fees from Pfizer, GE Health Care, Astra Zeneca, Novo Nordisk, investigator-initiated grant from Pfizer, GE Healthcare, Attralus, Siemens, Philips.

The other authors do not have any conflicts of interest to declare.

## Data availability statement

The data underlying this article cannot be shared publicly due to data privacy as defined in the informed consent document.

## Graphical Abstract

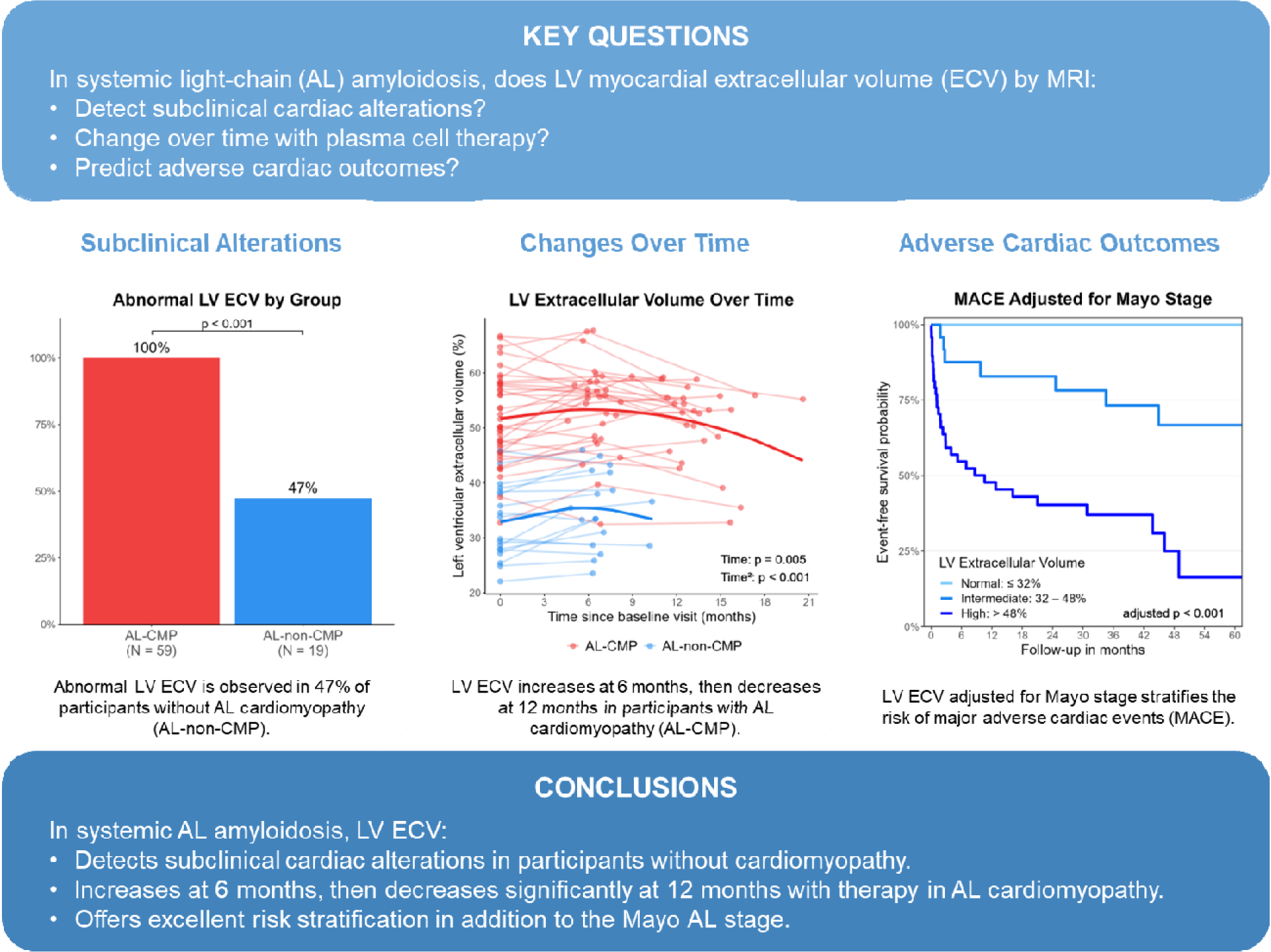

